# Ethics practices associated with reusing health data: An assessment of patient registries

**DOI:** 10.1101/2024.04.26.24306459

**Authors:** Olmo R. van den Akker, Susanne Stark, Daniel Strech

## Abstract

**Background:** As routinely collected patient data have become increasingly accessible over the years, more and more attention has been directed at the ethics of using such data for research purposes. Patient data is often available to researchers through patient registries that typically collect data of patients with a specific disease. While ethical guidelines for using patient data are presented frequently in research papers and institutional documents, it is currently unknown how patient registries implement the recommendations from these guidelines in practice and how they communicate their practices. In this project, we assessed to what extent a sample of 51 patient registries provides information about a range of ethics practices.

**Methods:** We searched for patient registries in the resource database of the European Network of Centres for Pharmacoepidemiology and Pharmacovigilance (ENCePP). Our assessment checklist was based on three sources: REQueST, a tool for the assessment of registry quality, AHRQ’s guide for good registry practices, and a systematic review of the principles and norms related to health data sharing by Kalkman and colleagues. The checklist includes 26 questions about five ethics components: governance, conflicts of interest, informed consent, privacy, and use-and-access.

**Results:** We found substantial heterogeneity in the way patient registries provide information about ethics practices: some registries rarely provide information; others discuss all relevant practices and more. Patient registries often mentioned their governance structure and any potential conflicts of interests but typically did not describe the responsibilities and rights allocated to their funders. Information about informed consent was often provided to patients but the available documents often lacked relevant information like the benefits and risks of participation. Privacy and use-and-access policies were typically discussed but not very concrete.

**Conclusions:** We can conclude that registries typically provide information about key ethics practices such as governance, conflicts of interest, informed consent, privacy, and use-and-access procedures, but that this information is often not as detailed as recommended in existing guidelines. The checklist we designed for our assessment could be helpful for the ethical assessments of patient registries and other types of registries in the future, as well as for self-assessment of registries that aim to improve their ethics practices.

## Background

As routinely collected patient data has become increasingly accessible for secondary use in research, regulatory practices, and health care, more and more attention has been directed at the ethics surrounding the use of such data. In a systematic review of the ethical principles and norms related to health data sharing Kalkman et al. (2019) identified four overarching themes: (1) societal benefits and value; (2) distribution of risks, benefits, and burdens; (3) respect for individuals and groups; and (4) public trust and engagement. These themes are echoed in two more formal guidelines that focus specifically on the data from patient registries (as opposed to health data in general). These registries are the main source for researchers to access patient data aside from clinical trials, giving them an important role in health research. First, the European Network for Health Technology Assessment (EUnetHTA, 2019) developed the Registry Evaluation and Quality Standards Tool (REQueST) to support more systematic and wide-spread use of registry data for regulatory purposes and health technology assessment (HTA). Even though they list ‘ethics’ as an additional instead of an essential requirement, they also include ethics practices like informed consent, governance, and data protection in their list of essential requirements. Second, the Agency for Healthcare Research and Quality (AHRQ, 2020) drafted a guide for good registry practices that includes a chapter in which they apply the Belmont principles of justice, beneficence, and respect for persons (National Commission for the Protection of Human Subjects of Biomedical and Behavioral Research, 1979) to the procedures of patient registries.

In sum, guidance is available on how to ethically share and use patient data. However, to implement broadly formulated ethical principles, a certain translation into concrete normative practices is needed (Schwietering, Langhof, & Strech, 2023). For example, informed consent reflects a normative practice that implements the principles ‘respect for individuals’ and ‘public trust’. Similarly, a clear governance structure with patient engagement reflects the ethical principles ‘public trust’ and ‘engagement’. It is currently unknown how patient registries implement the available ethics guidance in practice.

A review on the ethics in biobank research showed that only very few studies investigated the implementation of specific normative practices for secondary-use studies (Langhof, Schwietering, & Strech, 2018). The authors of this review developed a matrix of nine normative practices with relevance to biobank research: informed consent, independent ethics review, data safety and security, sample ownership, sample access and compensation, priority setting, incidental findings, public and patient involvement, and ethics reporting. This review, however, assessed the *normative* research literature on biobank ethics, not the way that biobanks *actually* handle the normative practices. Pavlenko, Strech, and Langhof (2020) did directly assess how often and to what extent data warehouses make their use-and-access policies publicly available and what selection criteria they contained, but they focused only on that specific practice.

In this paper, we aimed to assess to what extent a sample of patient registries provides information about a wide range of ethics practices. The results of this project shed light on how patient registries report about their current ethics practices and will highlight where improvements can be made.

## Methodology

### Sample selection

We searched for patient registries in the resource database of the European Network of Centres for Pharmacoepidemiology and Pharmacovigilance (ENCePP). ENCePP is a network coordinated by the European Medicines Agency (EMA) that involves public institutions and organizations involved in pharmacoepidemiology and pharmacovigilance research. We chose this registry because our project is part of an EU project (More-EUROPA) on secondary use of patient data within the context of regulatory decision making and HTA. In consultation with other members of More-EUROPA we searched for an established database listing patient registries with relevance for regulatory and HTA activities. The ENCePP database was decided on unanimously. In the ENCePP resource database we searched for types of data sources that we believed would include patient registries: ‘Disease/case registry’, ‘Routine primary care electronic patient registry’, ‘Exposure registry’, and ‘Other’. We further limited our search by only including registries from the European Economic Area, because of its relevance to the More-EUROPA project. This search strategy is in line with Plueschke, Jonker, Strassmann, & Kurz (2022) who targeted patient registries to conduct a survey to better understand their approach towards the collection, management and reporting of adverse effects related to medicines. For feasibility purposes we excluded transnational registries leading to a sample of 68 registries (see Table 1 for an overview). This search was conducted before coding and after preregistering our study on the Open Science Framework (see https://osf.io/rdfs2). During coding, we had to exclude 17 of these 68 registries, yielding a total sample of 51 registries for our assessment (see the Results section for more details about these exclusions).

**Table 1.**
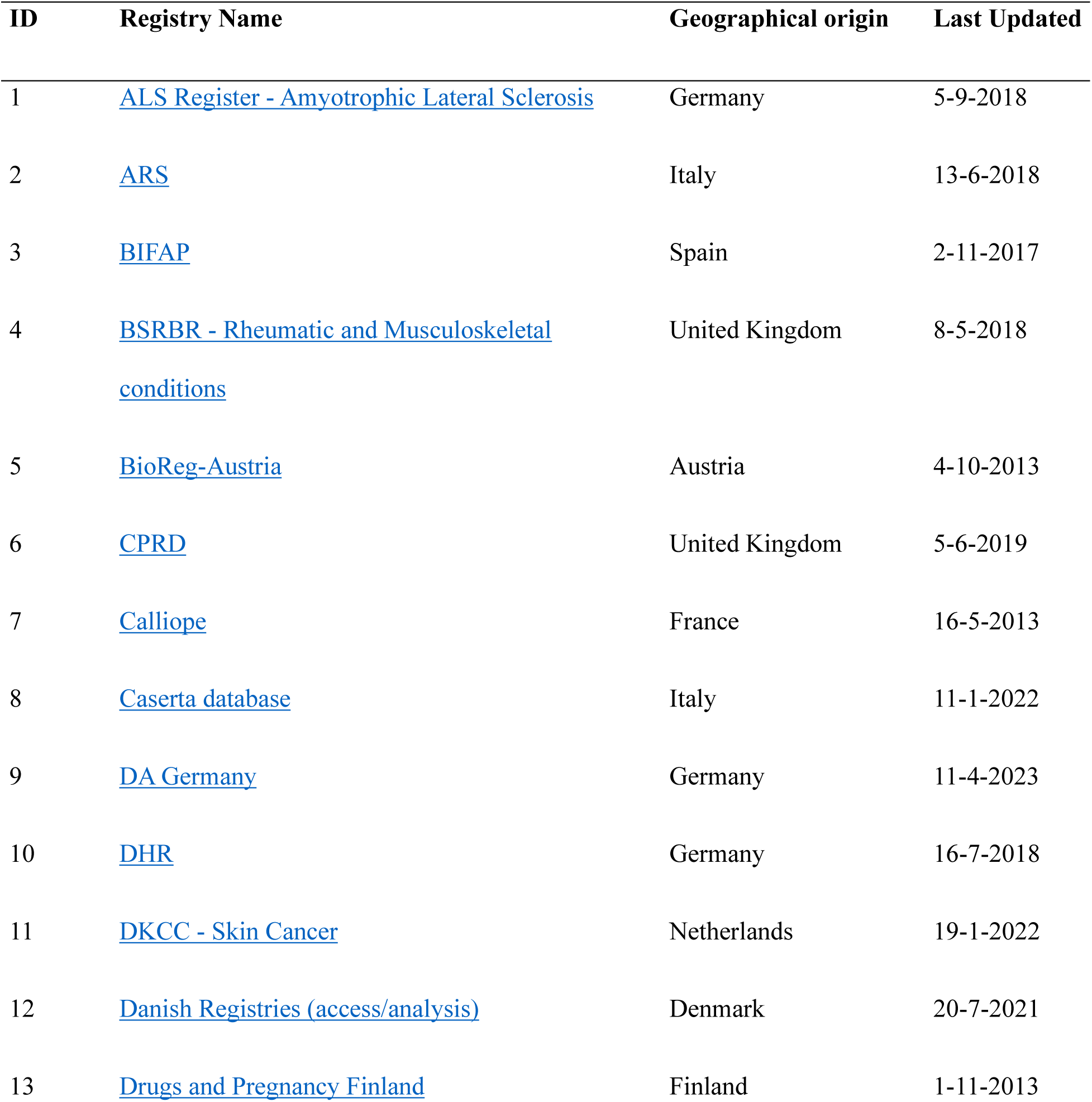
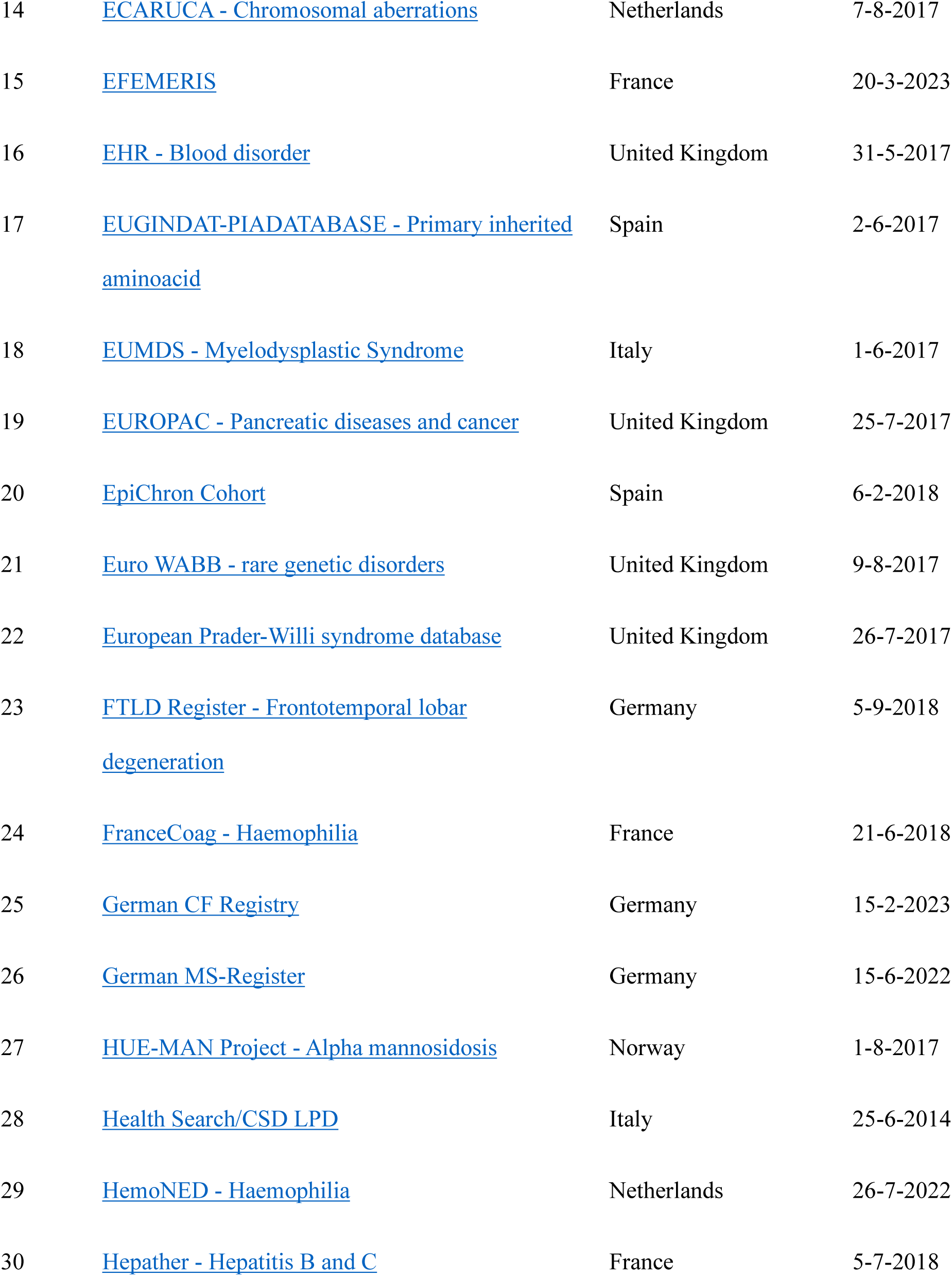
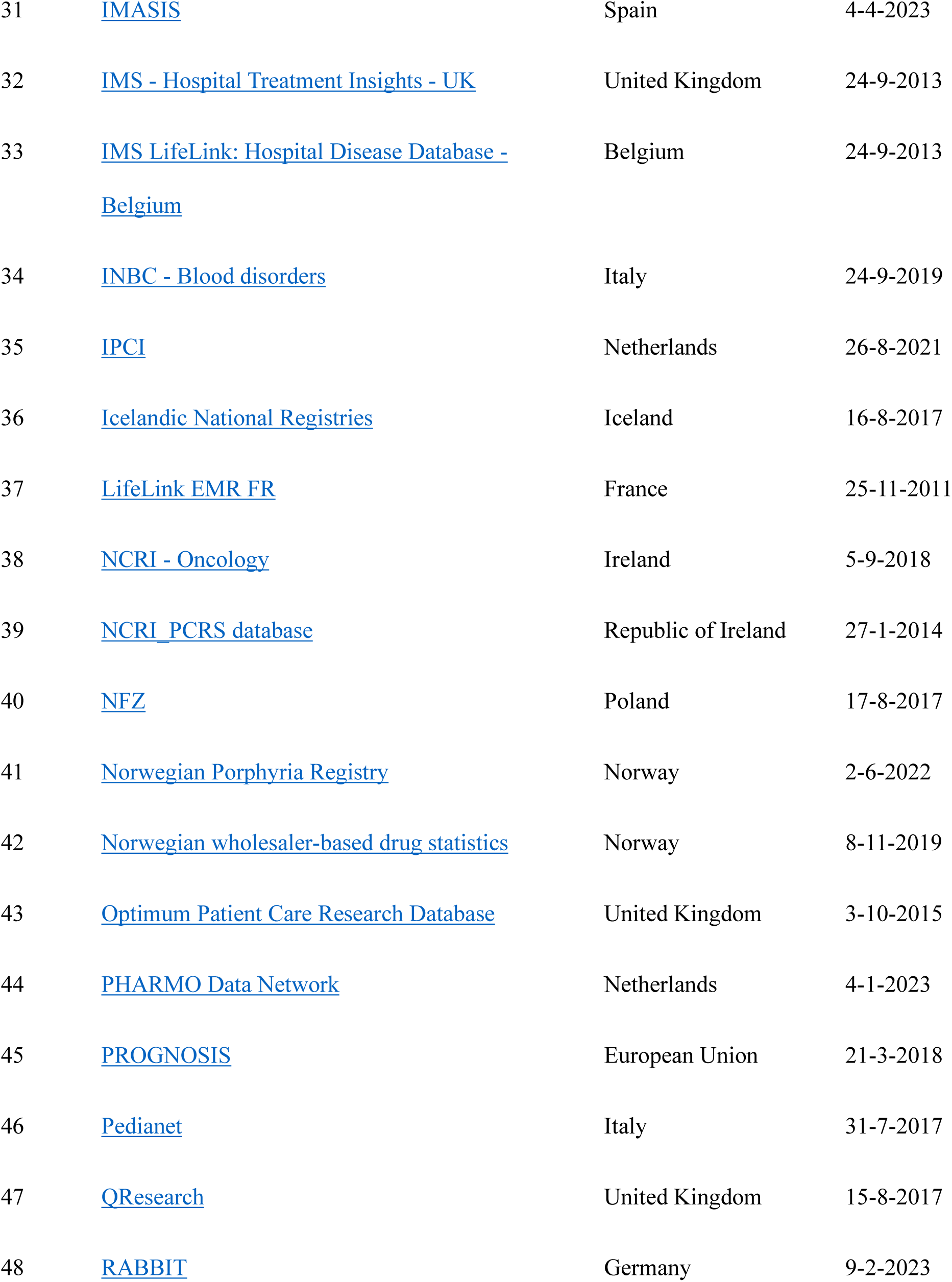
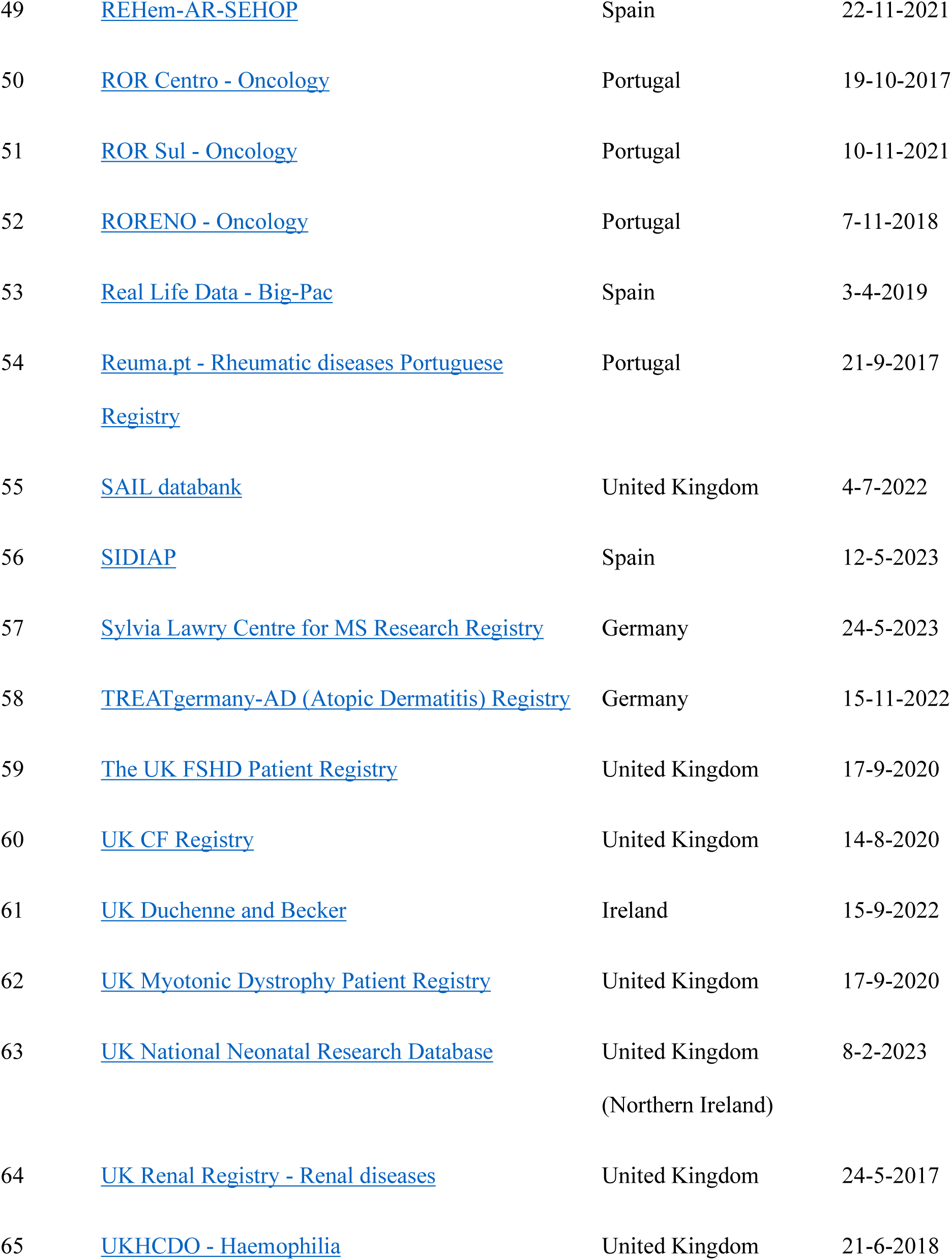
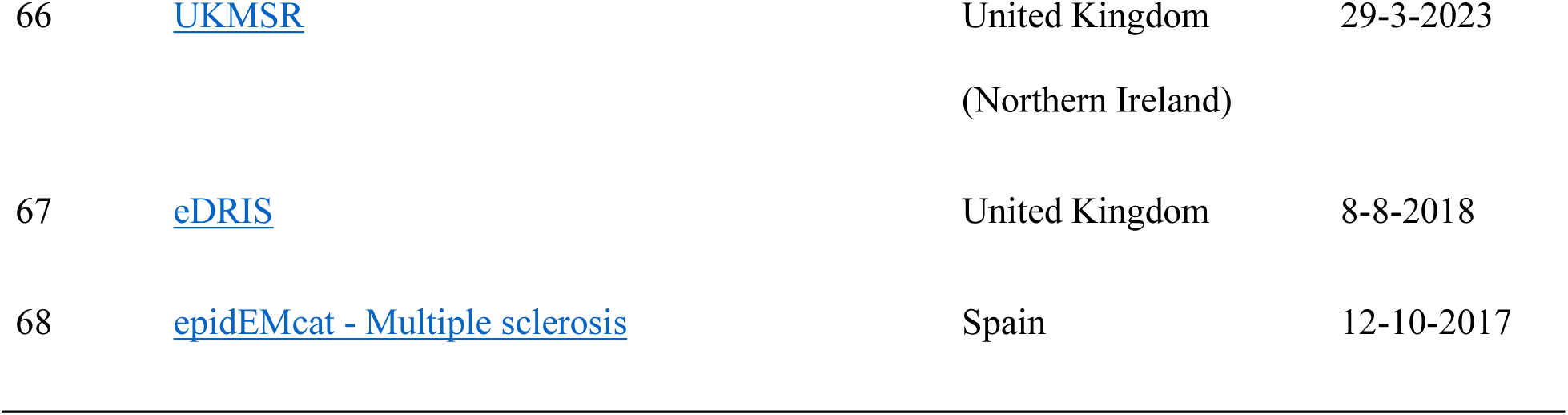
Patient registries that were assessed in this study.

### Assessment checklist

As mentioned in our preregistration (https://osf.io/rdfs2) the starting point for the development of our ethics checklist was REQueST, a tool for the assessment of registry quality (EUnetHTA, 2019). This tool involves a list of twelve essential and three additional requirements that high-quality registries should fulfil. From this list, we identified five requirements related to ethics: Governance; Informed consent; Financing; Protection, security and safeguards; and Ethics. Table 2 provides more information about each of these requirements.

**Table 2.**
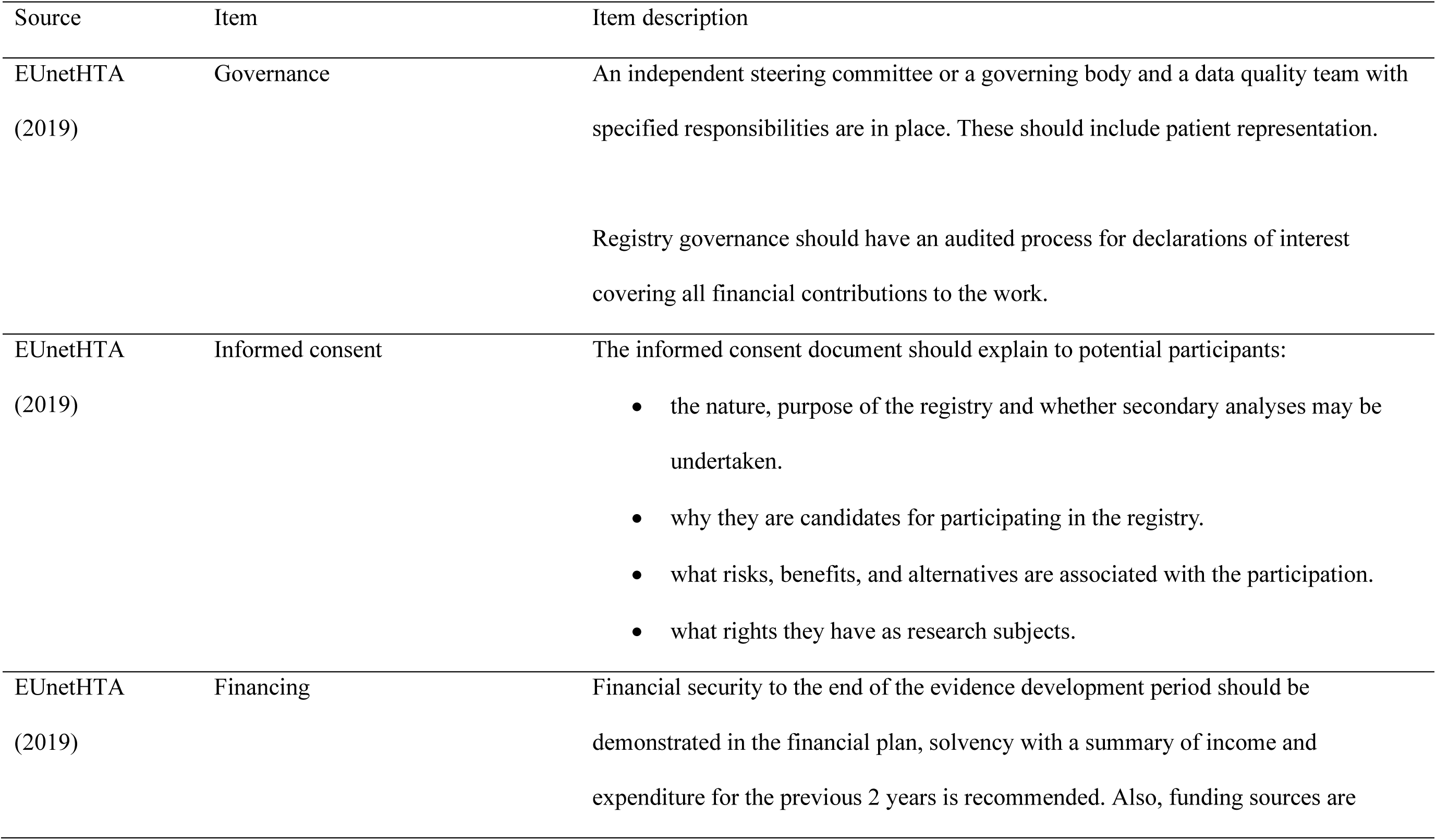
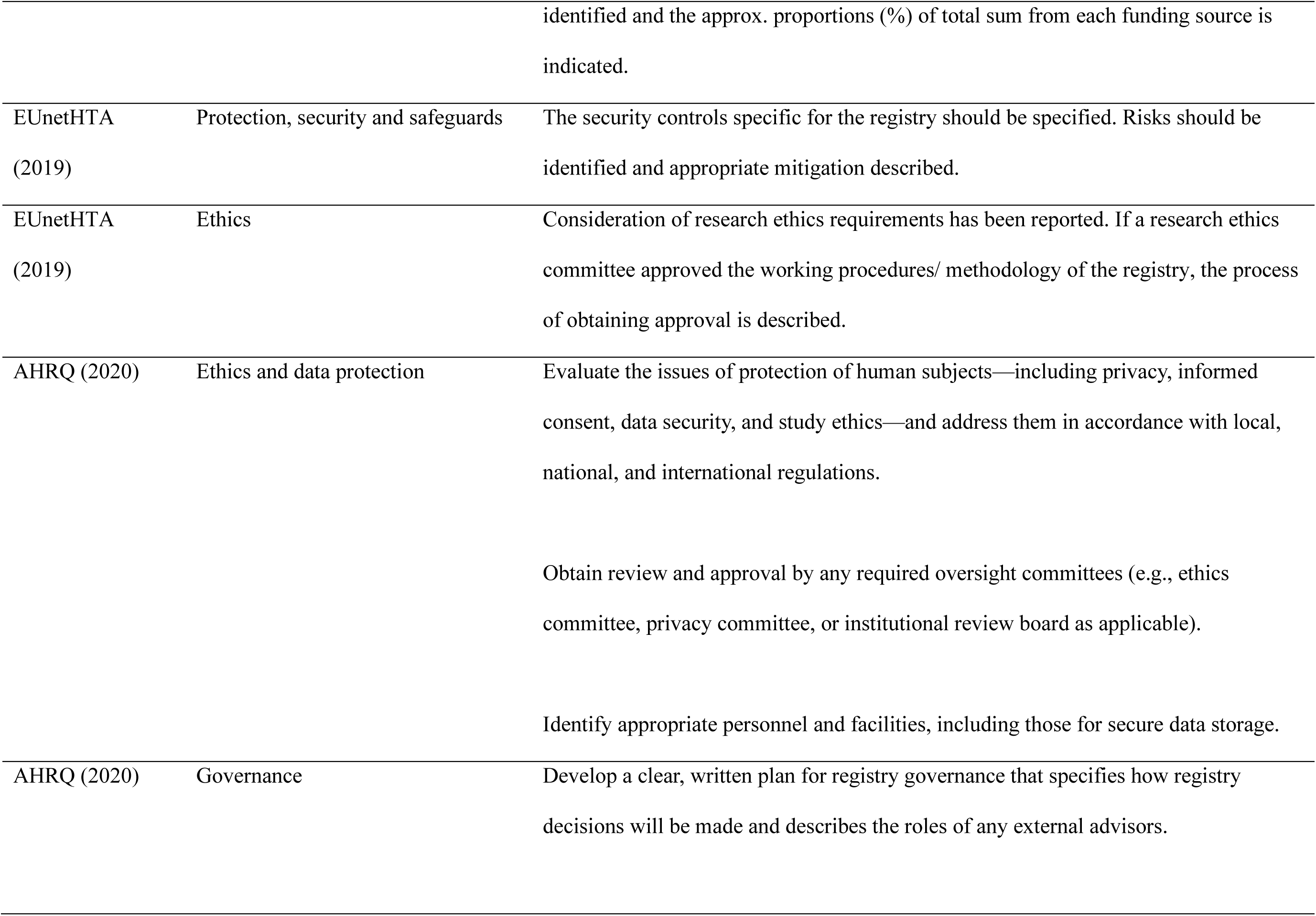
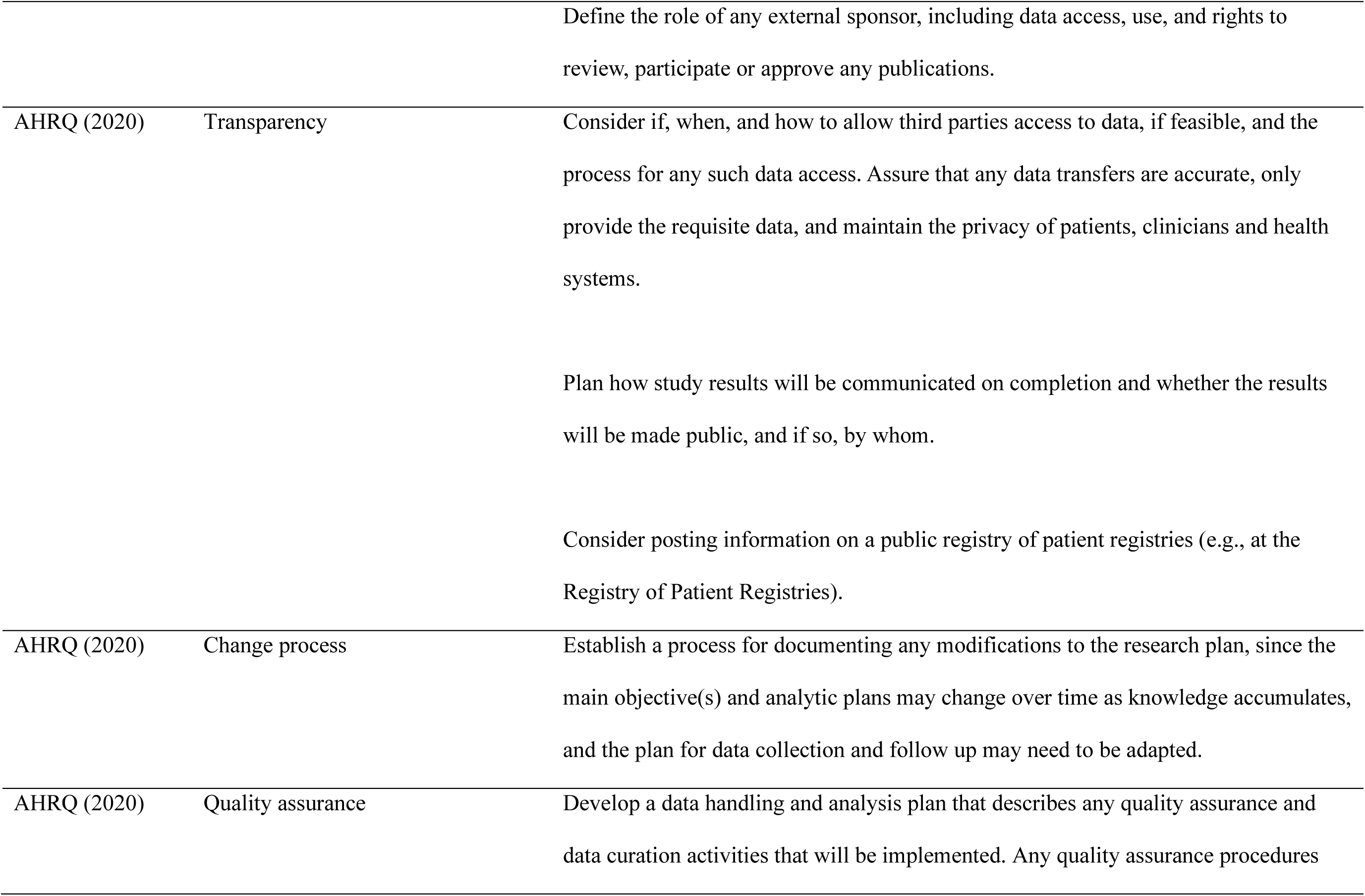
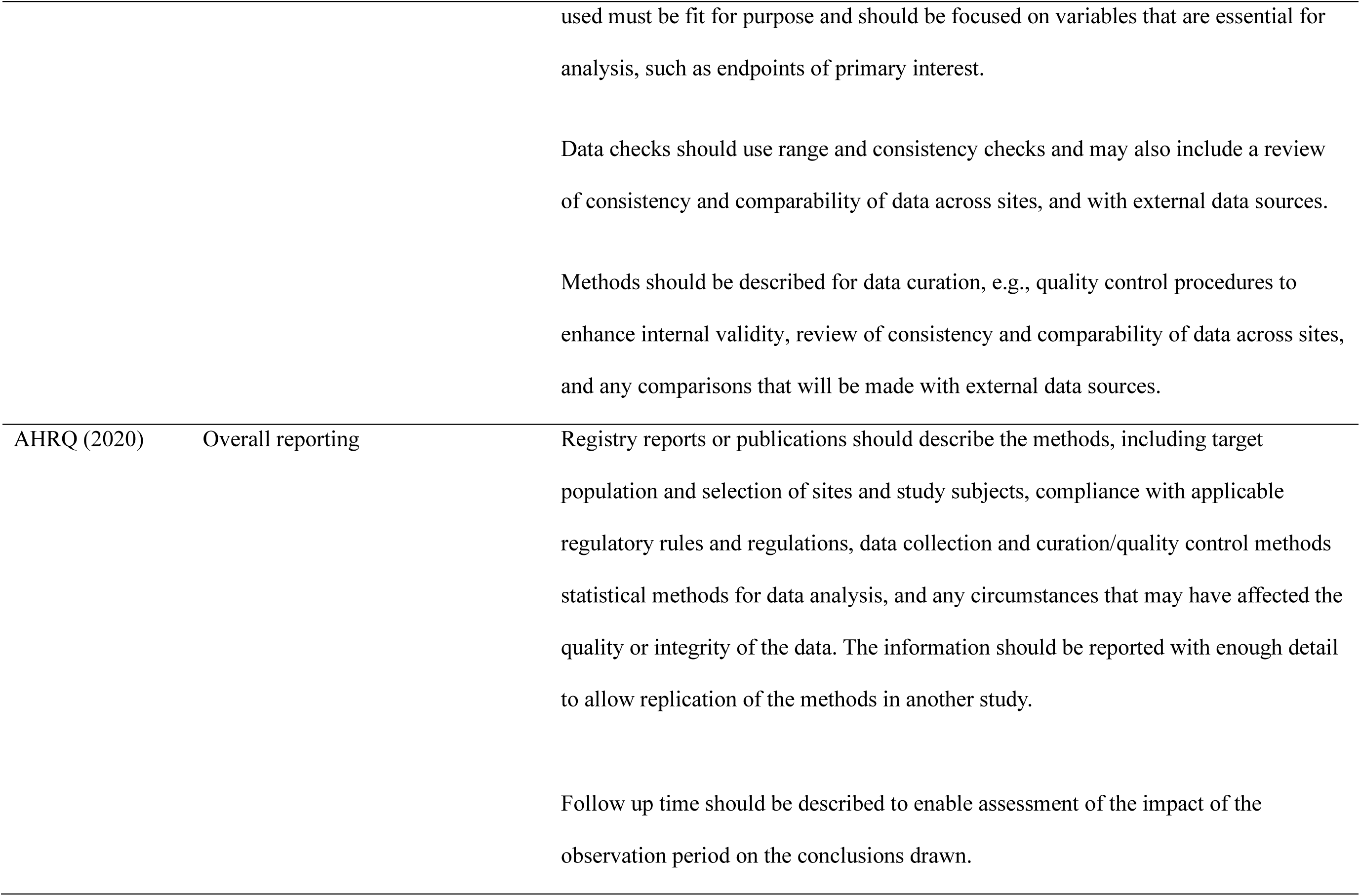
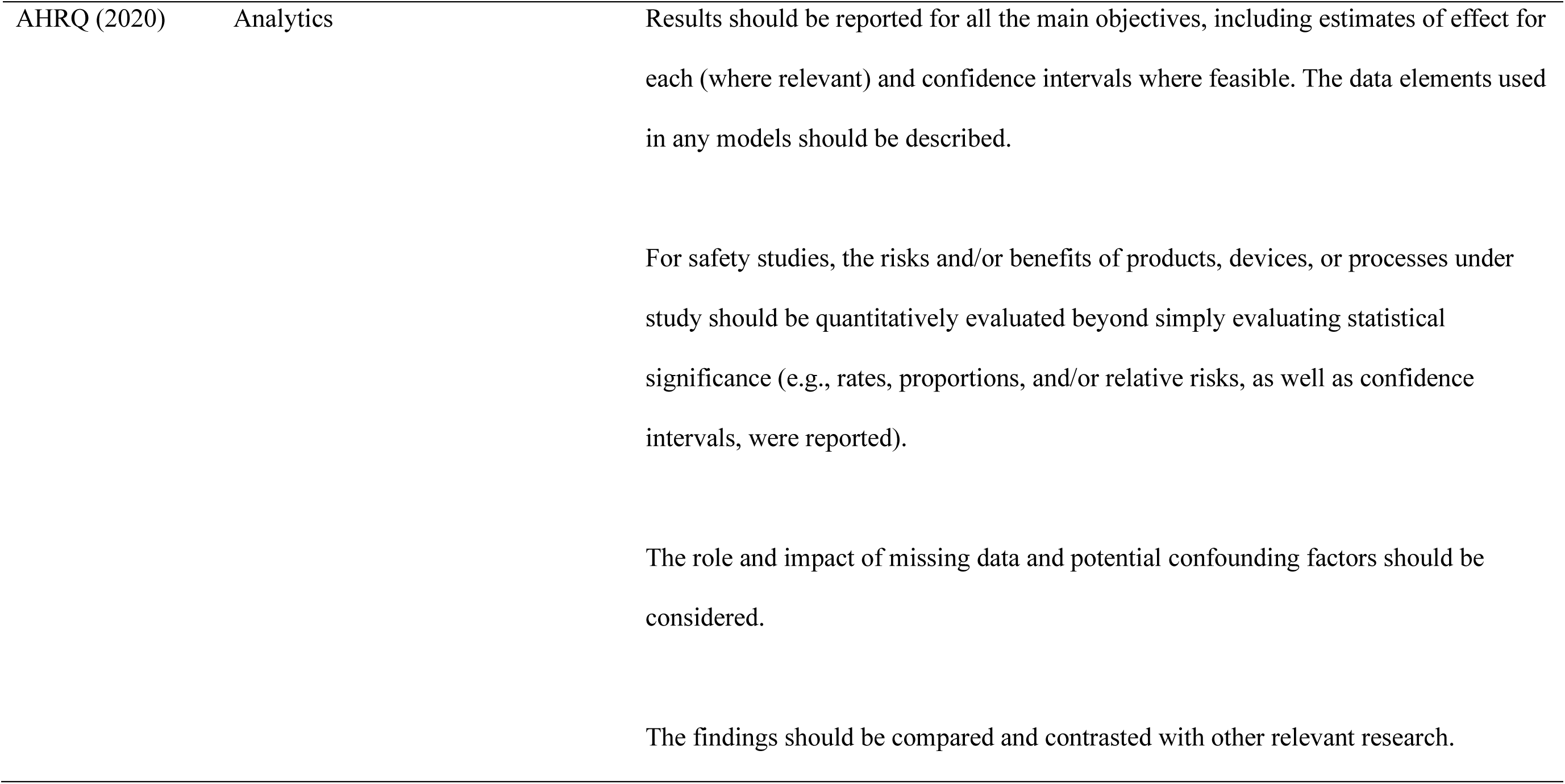
Items used as background for our assessment checklist.

Even though the REQueST tool captures important ethics practices, there were some practices missing based on the principles provided by Kalkman et al. (2019). Therefore, we decided to extend the list of requirements with several items from AHRQ’s (2020) guide for good registry practice. This guide serves as “*a reference for planning, developing, maintaining, and evaluating registries designed to collect data about patient outcomes*” and is therefore well-suited to our goals. The items we identified as relevant were part of the domain categories ‘Framework’ (Ethics and data protection; Governance; Transparency; and Change process), ‘Methods’ (Quality assurance), and ‘Reporting’ (Overall reporting; and Analytics). Table 2 provides more details about these items. Note that the description of these items is written from the perspective of a registry owner as that is the guide’s target audience.

As an overarching third source we used Kalkman et al.’s (2019) systematic review of the principles and norms related to health data sharing. In their review, the authors do not provide concrete items to assess registries but do mention several themes and principles relevant to ethics practices in the context of the secondary use of health data, like informed consent, privacy, and use-and-access. We used these topics to make our checklist more fine-grained. For example, the review stressed that use-and-access policies should include a procedure to prevent unauthorized access and should explicitly state that data users must refrain from any attempt to (re-)identify patients. We updated our checklist by adding items to assess these practices.

Finally, we used our own expertise to synthesize the information from the three sources into concrete items suitable for our checklist. This synthesis mainly consisted of extracting phrases related to the aforementioned ethics practices, merging phrases from the different sources, and transforming each item into a question. We then piloted the checklist by coding the information provided on the websites of the first ten registries listed in Table 1. This was done by two independent coders (OvdA and SS) in frequent consultation with DS. Any coding complexities identified in the coding were solved through a group discussion of the author team. The aim of the pilot was to weed out any ambiguity or unclarity in the phrasing of the questions, and to ensure that there would be no idiosyncrasies in the coding of the rest of the registries. The rest of the registries were coded by OvdA alone. A practice received a code of 1 when information was provided about it on the website and a code of 0 when no information was provided. Any exceptions to this binary classification can be found in Table 3, which provides the final checklist used in our coding. Note that this final checklist deviates somewhat from the checklist we preregistered (see https://osf.io/rdfs2 for the preregistration and https://osf.io/b2uj4 for the list of changes).

**Table 3.**
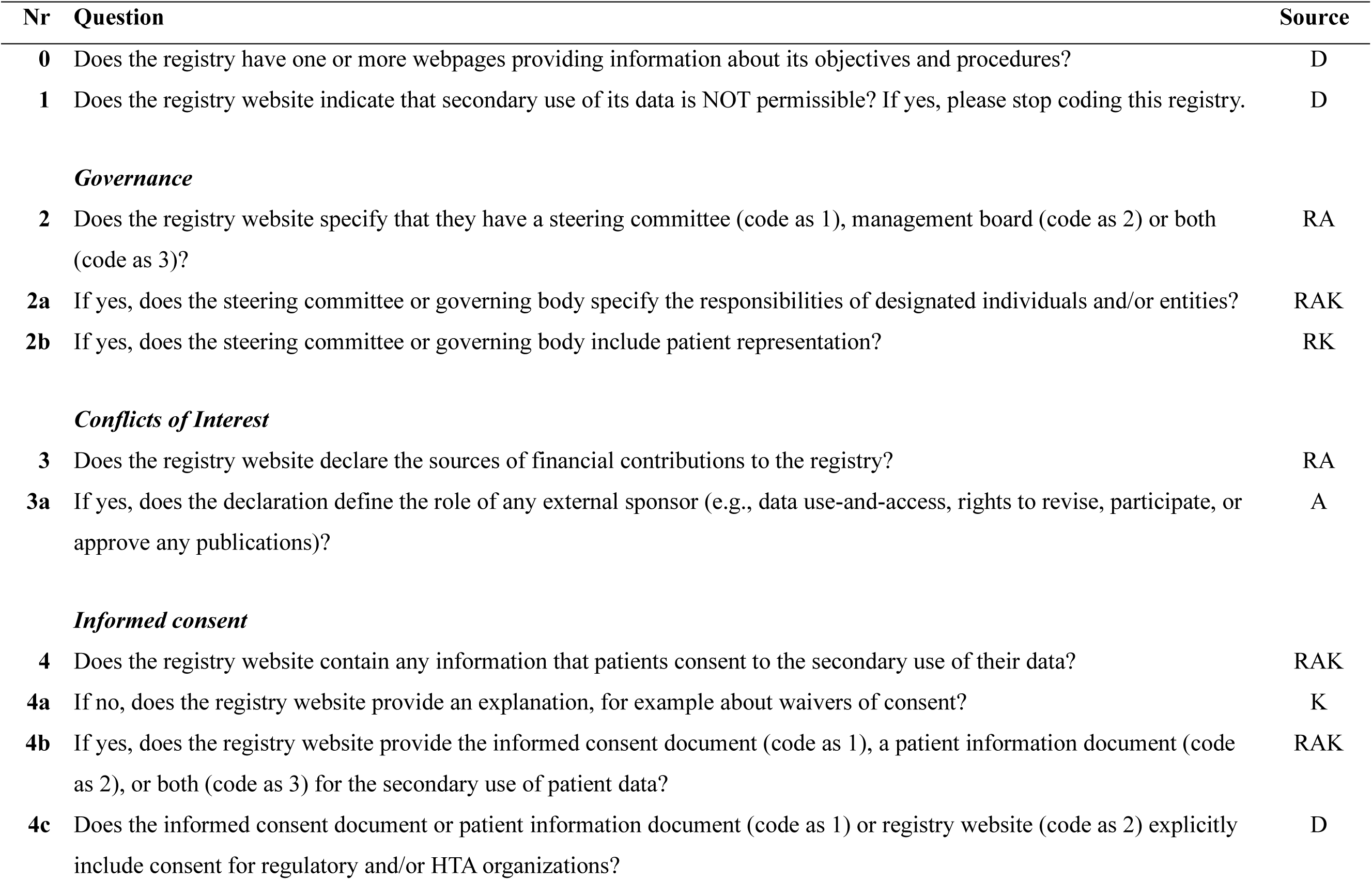
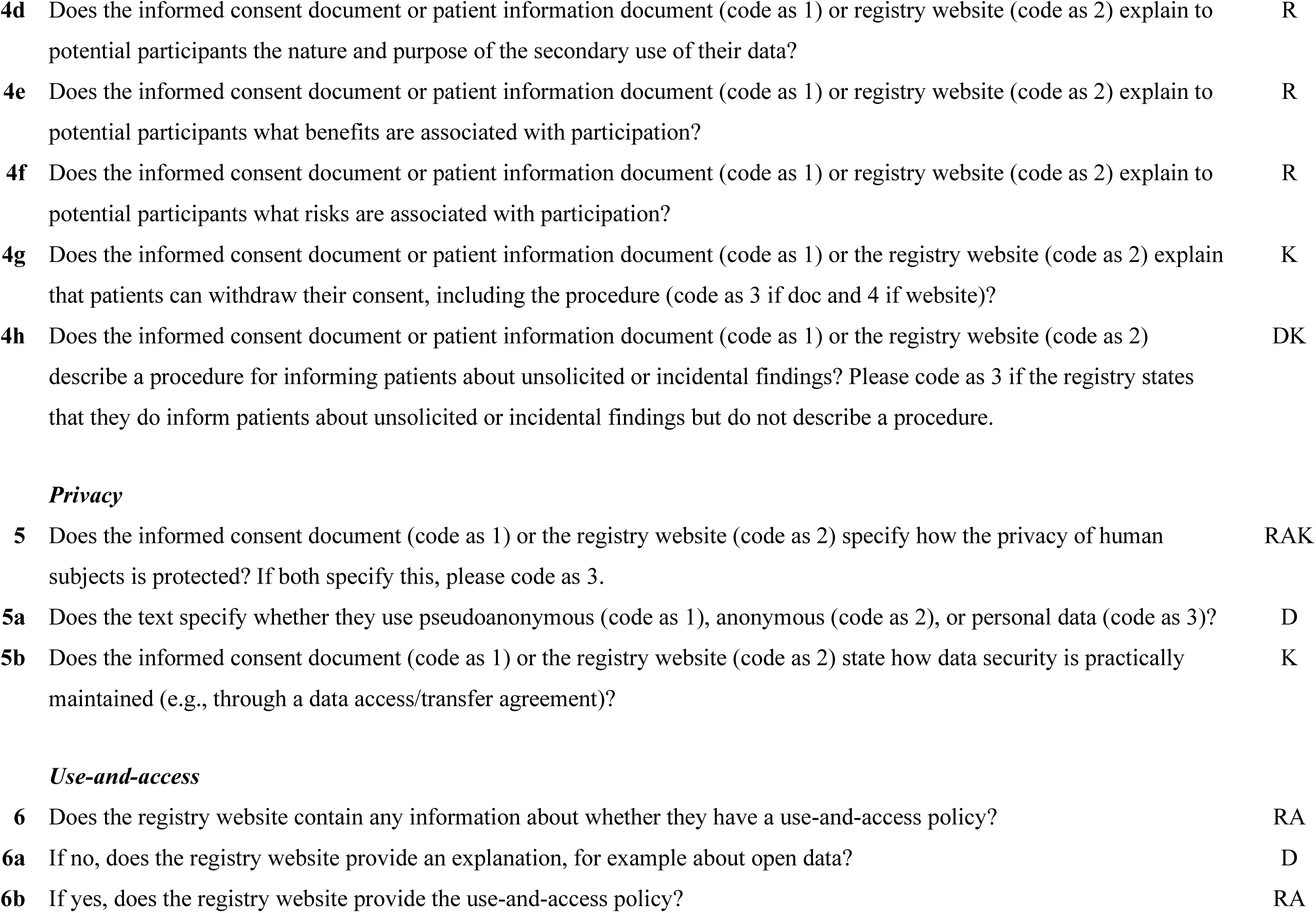
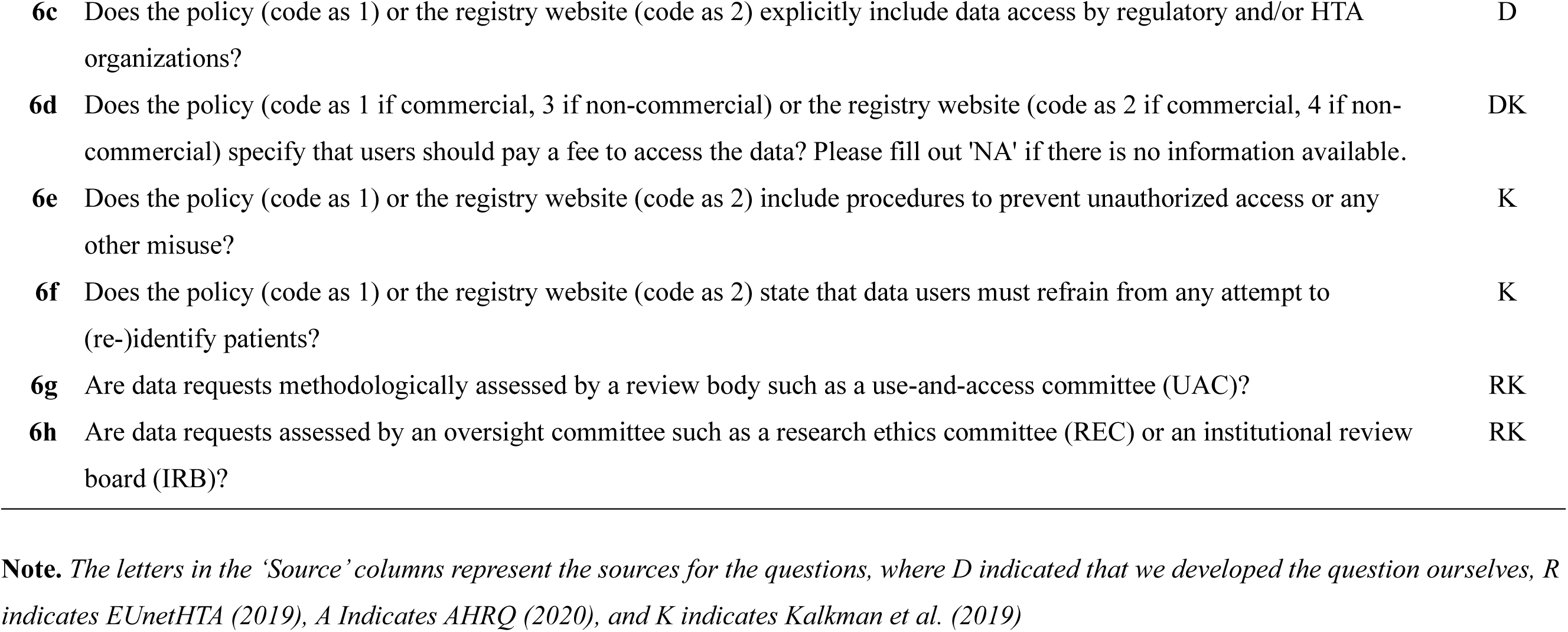
The checklist questions used to assess the ethics of patient registries.

| Nr | Question | Source |
| --- | --- | --- |
| 0 | Does the registry have one or more webpages providing information about its objectives and procedures? | D |
| 1 | Does the registry website indicate that secondary use of its data is NOT permissible? If yes, please stop coding this registry. | D |
| ***Governance*** |  |  |
| 2 | Does the registry website specify that they have a steering committee (code as 1), management board (code as 2) or both (code as 3)? | RA |
| 2a | If yes, does the steering committee or governing body specify the responsibilities of designated individuals and/or entities? | RAK |
| 2b | If yes, does the steering committee or governing body include patient representation? | RK |
| ***Conflicts of Interest*** |  |  |
| 3 | Does the registry website declare the sources of financial contributions to the registry? | RA |
| 3a | If yes, does the declaration define the role of any external sponsor (e.g., data use-and-access, rights to revise, participate, or approve any publications)? | A |
| ***Informed consent*** |  |  |
| 4 | Does the registry website contain any information that patients consent to the secondary use of their data? | RAK |
| 4a | If no, does the registry website provide an explanation, for example about waivers of consent? | K |
| 4b | If yes, does the registry website provide the informed consent document (code as 1), a patient information document (code as 2), or both (code as 3) for the secondary use of patient data? | RAK |
| 4c | Does the informed consent document or patient information document (code as 1) or registry website (code as 2) explicitly include consent for regulatory and/or HTA organizations? | D |
| **4d** | Does the informed consent document or patient information document (code as 1) or registry website (code as 2) explain to potential participants the nature and purpose of the secondary use of their data? | R |
| **4e** | Does the informed consent document or patient information document (code as 1) or registry website (code as 2) explain to potential participants what benefits are associated with participation? | R |
| **4f** | Does the informed consent document or patient information document (code as 1) or registry website (code as 2) explain to potential participants what risks are associated with participation? | R |
| **4g** | Does the informed consent document or patient information document (code as 1) or the registry website (code as 2) explain that patients can withdraw their consent, including the procedure (code as 3 if doc and 4 if website)? | K |
| **4h** | Does the informed consent document or patient information document (code as 1) or the registry website (code as 2) describe a procedure for informing patients about unsolicited or incidental findings? Please code as 3 if the registry states that they do inform patients about unsolicited or incidental findings but do not describe a procedure. | DK |
| **5** | Does the informed consent document (code as 1) or the registry website (code as 2) specify how the privacy of human subjects is protected? If both specify this, please code as 3. | RAK |
| **5a** | Does the text specify whether they use pseudoanonymous (code as 1), anonymous (code as 2), or personal data (code as 3)? | D |
| **5b** | Does the informed consent document (code as 1) or the registry website (code as 2) state how data security is practically maintained (e.g., through a data access/transfer agreement)? | K |
| **6** | Does the registry website contain any information about whether they have a use-and-access policy? | RA |
| **6a** | If no, does the registry website provide an explanation, for example about open data? | D |
| **6b** | If yes, does the registry website provide the use-and-access policy? | RA |
| **6c** | Does the policy (code as 1) or the registry website (code as 2) explicitly include data access by regulatory and/or HTA organizations? | D |
| **6d** | Does the policy (code as 1 if commercial, 3 if non-commercial) or the registry website (code as 2 if commercial, 4 if non-commercial) specify that users should pay a fee to access the data? Please fill out 'NA' if there is no information available. | DK |
| **6e** | Does the policy (code as 1) or the registry website (code as 2) include procedures to prevent unauthorized access or any other misuse? | K |
| **6f** | Does the policy (code as 1) or the registry website (code as 2) state that data users must refrain from any attempt to (re-)identify patients? | K |
| **6g** | Are data requests methodologically assessed by a review body such as a use-and-access committee (UAC)? | RK |
| **6h** | Are data requests assessed by an oversight committee such as a research ethics committee (REC) or an institutional review board (IRB)? | RK |

**Note.** The letters in the ‘Source’ columns represent the sources for the questions, where D indicated that we developed the question ourselves, R indicates EUnetHTA (2019), A Indicates AHRQ (2020), and K indicates Kalkman et al. (2019)

## Results

For each of the 68 registries in our sample, we first tried to access the website listed in the ENCePP entry. A substantial number of websites (n = 11) were no longer functioning, possibly because the registries were discontinued (e.g., the Caserta database and the HUE-MAN patient registry on alpha mannosidosis). At the same time, two ENCePP links did not lead to a website with information about the registry itself but instead to a website with information about the organization hosting the registry (i.e. the ARS registry with http://www.ars.toscana.it, and the European Haemoglobinopathy Registry with http://www.sickle-thal.nwlh.nhs.uk). Moreover, it appeared that some registries changed their name (e.g. BSRBR - Rheumatic and Musculoskeletal Conditions to Rheumatoid Arthritis Register) or were subsumed into a larger, often commercial, registry collective (e.g. DA Germany and several IMS Health registries were transformed into IQVIA). Changes like these complicated our efforts to find an official website for each individual registry. In total, we were able to retrieve informative websites for 56 registries. Out of these 56 registries, three did not have procedures in place to make their data available to external researchers (i.e. the European registry of hereditary pancreatitis and familial pancreatic cancer, EpiChron Cohort, and the Scottish Electronic Data Research and Innovation Service). Instead, the data could only be used by a pre-specified internal group or data access was not yet enabled. Excluding these three registries and two duplicates (the Frontotemporal lobar degeneration Register was already coded under the name ALS registry, while the National Cancer Registry Ireland had two entries in ENCePP), our final sample consisted of 51 registries. Our findings regarding these registries are presented per ethics theme below. Table 4 provides the frequencies of all coded responses. An overview of the raw data including the source and explanation for each individual code can be found at https://osf.io/wmrv2. The code we used to summarize the data can be found at https://osf.io/twfu6.

**Table 4.**
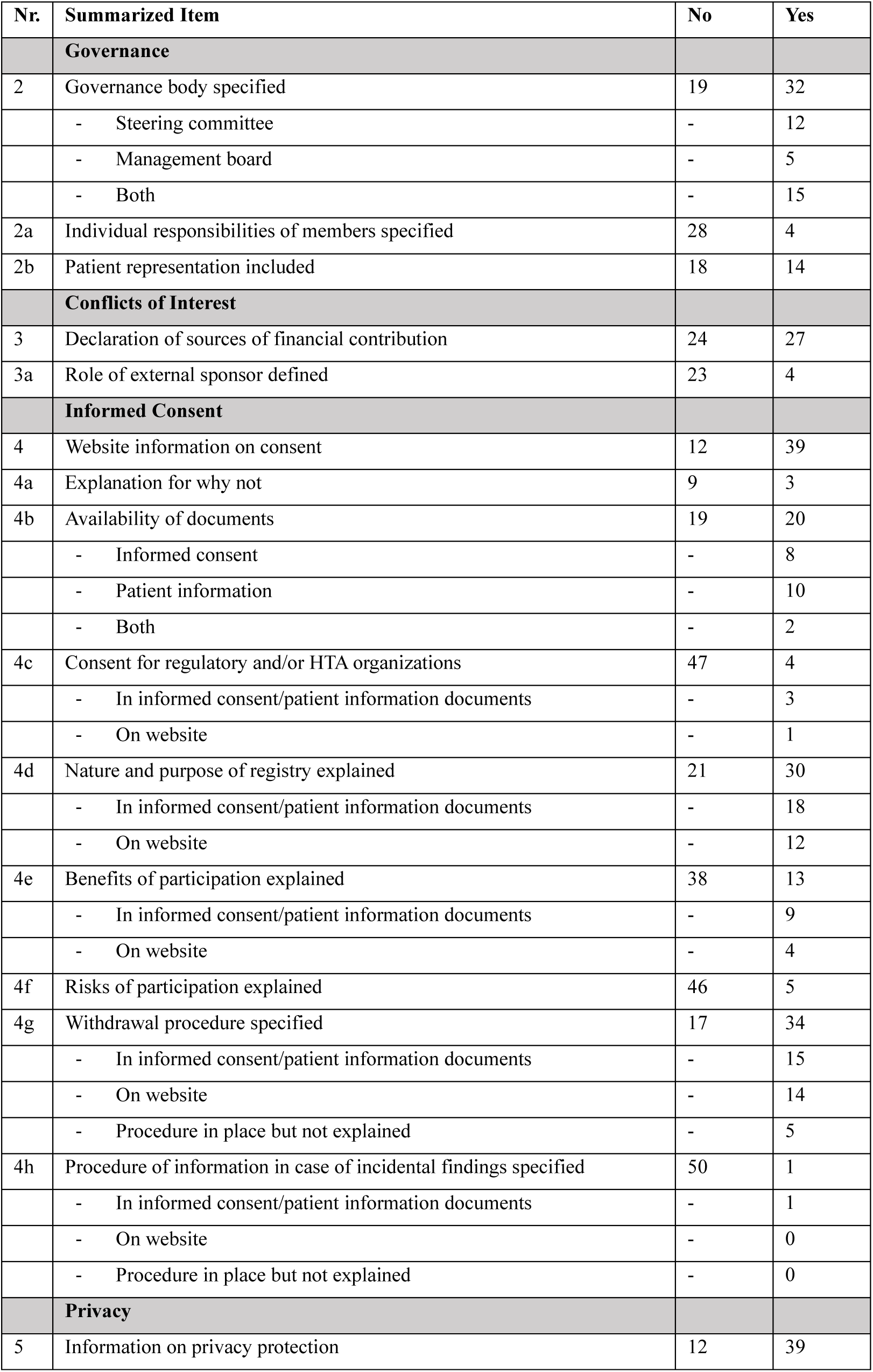
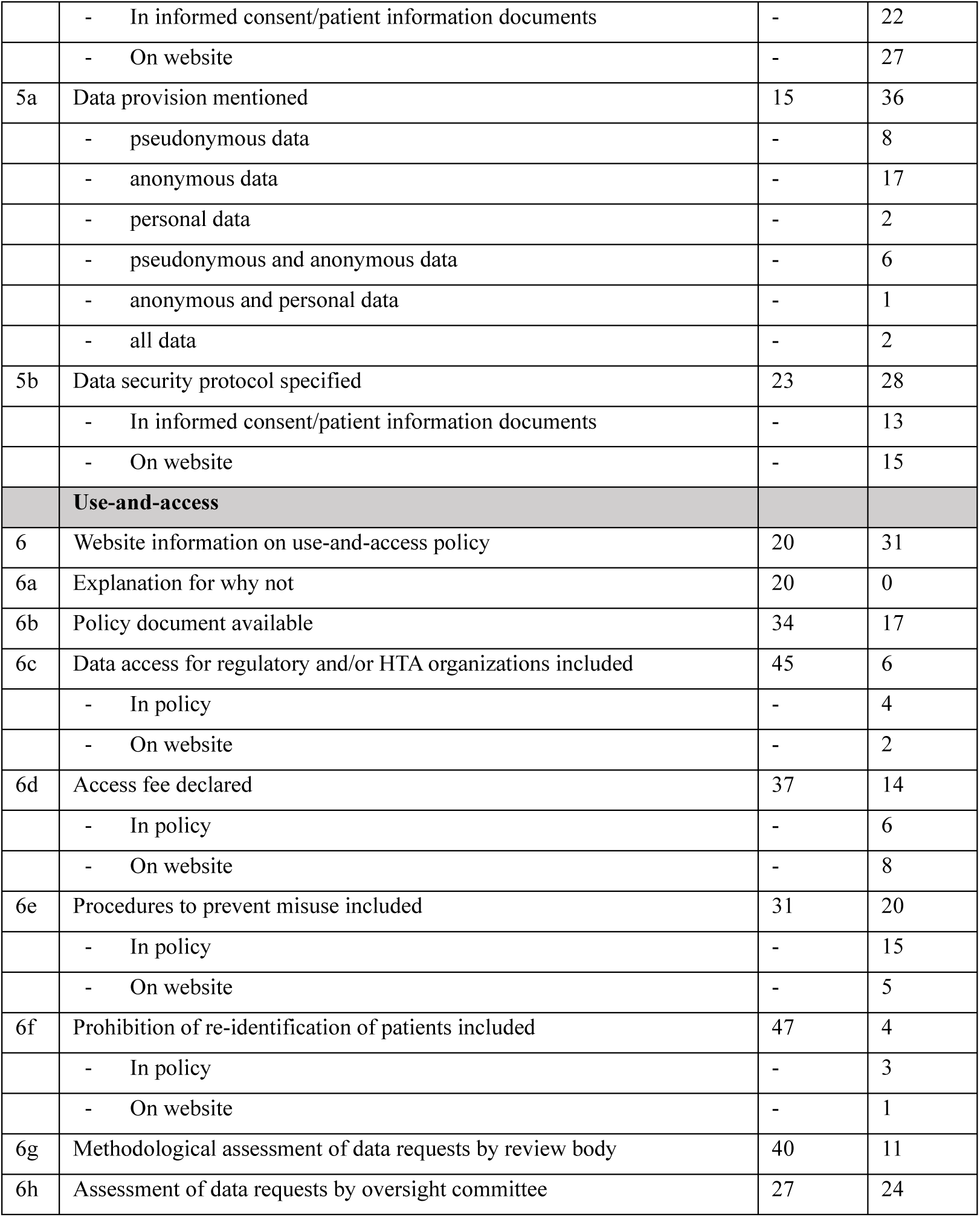
The coded responses for all checklist questions.

### Governance and Conflicts of interest

Out of the 51 coded registries, 19 (37%) did not provide information about having a governing body (responsible for the day-to-day management of the registry) or a steering committee (to supervise the daily management of the registry) on their website. Twelve registries (24%) reported to have a steering committee, five (10%) only stated to have a governing body, and fifteen (29%) reported to have both. Importantly, of the 32 registries reporting some form of governance, only 4 (13%) provided the tasks and responsibilities of individual members of the steering committee / governing body. Furthermore, fourteen of these 32 registries (44%) included patient representation in their steering committee or governing body. That being said, some registries that did not report formal patient representation did have other ways of involving patients in their procedures. For example, the UK Renal Registry has an active patient council that aims to solicit experiences and opinions from patients about their services, while the UK Multiple Sclerosis Register has a dedicated online group of participants who provide feedback on any new features, ideas, and questionnaires.

Twenty-seven out of the 51 registries (53%) mentioned on their website how the registry is financed. However, only three of those (11%) also provided details about the influence these financiers have on the registry procedures.

### Informed consent and Privacy

Most registries (n = 39, 76%) mentioned the concept of informed consent on their website, but only about half of those (n = 20, 51%) provided the informed consent form itself (n = 8, 21%), a patient information document (n = 10, 26%), or both (n = 2, 5%). Of the registries that did not mention that informed consent was relevant (n = 12, 24%), three provided an explanation of why that was the case. In all cases that was because they only use anonymized data. The other nine registry websites (18%) did not mention anything about informed consent at all.

We also assessed the information that was provided in the consent forms and patient information documents. In this analysis, we bundled these two types of forms together because they involve broadly the same informative function, leading to 20 information forms. The three sources we used to develop the checklist provided several topics that are important to include in forms like this. Eighteen forms (90%) included information about the aims of the registry. While the benefits of participating in the registry were included relatively often (n = 13, 65%), the risks were rarely mentioned (n = 5, 25%). The most mentioned benefit for patients was the improvement of their own health as well as those of patients with the same disease. In paragraphs about potential risks, registries often mentioned that there were none. All the 20 documents specified that participants have a right to withdraw their consent, although five of them (25%) were not clear about the procedure patients would need to follow to set such a withdrawal in motion. Finally, only one of the documents (5%) indicated a procedure to inform patients about incidental findings (i.e., that of the European Registry for Myelodysplastic Syndromes). The registries that mentioned consent but did not provide information documents (n = 19, 49%) did typically provide information elsewhere on their website. Twelve (63%) provided information about the nature and purpose of the registry, four (21%) provided information about the benefits of participation, fourteen (74%) provided information about how to withdraw consent, and none provided information about the risks of participation or incidental findings. We do not know whether patients were actually presented the information on the website when they were asked to contribute their data.

While privacy can be seen as an important part of informed consent forms, during coding we decided to include it as a separate category given that many registry websites dedicate separate sections to this topic as opposed to including it in informed consent or patient information documents. Most registries provided some information on how patient privacy is handled (n = 39, 76%). The amount of information provided differed substantially, however. Some registries merely mentioned that privacy was ensured (e.g. the IQVIA registries and the Italian National Blood Centre), while others provided extensive privacy policies including many different perspectives and even explanations about different components of privacy (e.g. the German Cystic Fibrosis Registry and the National Cancer Registry Ireland). Most registries that mentioned something about privacy (n = 28, 72%) dedicated one or more paragraphs to the topic of data security. Finally, seventeen (33%) of the registries in our sample stated that they used anonymous data, eight registries (16%) that they used pseudonymous data (i.e., data with an identifier that cannot be reduced to patients without a key that is stored with a trusted party), two registries (4%) that they used personal data, and nine registries (18%) used a combination (i.e., in six registries [12%] some data are pseudonymized and other data are anonymized; in one registry [2%] some data are anonymized and other data are personal; in two registries [4%] all three data types are present). The other fifteen registry websites (29%) did not mention this information explicitly.

### Use-and-access

The final practice we assessed was the use-and-access policies of registries. A majority of registries (n = 31) stated that they have a policy to decide who gets access to their data, but only seventeen of those provided a detailed page or document to outline the use-and-access criteria and procedure. In a typical procedure, interested researchers would fill out a submission form, either on the website directly, or using a document from the website that they could email later. This submission form often had to include a clear description of the research questions, the type of data required, and the statistical analysis planned (although this latter requirement was rarer). These submissions were then reviewed by a committee dedicated to this task by the registry. Often, registries did not go into detail about the specifics of this review, but five registry websites (35%) at least mentioned that it concerned a methodological review, 18 (77%) that it concerned an ethical review, and six (19%) that both an ethical and a methodological review took place. The ethics assessment could be done by either an ethics committee at the researchers’ institute or internally by an ethical committee or supervisory organ of the registry itself.

Of all registries that mentioned use-and-access (n = 31, 61%), twenty (65%) addressed how they would ensure that the registry data would not be misused by unauthorized people. Eight of the total registries in our sample (16%) stated that they require a fee from researchers to use their data, ten (20%) were free to use or required a small fee to ensure they would break even, and the others (n = 33, 65%) did not provide any information about fees. Finally, not many of the texts about use-and-access explicitly mentioned that researchers should refrain from trying to (re-)identify patients (n = 4, 31%).

## Discussion

In this project, we assessed to what extent patient registries provide information about their ethics practices. For this purpose, we examined a sample 68 patient registry websites from the ENCePP database using a tailor-made checklist. We found that the ENCePP database was not always up to date as we were able to find webpages with relevant information for only 51 registries. Based on that subsample, we can conclude that registries typically provide information about key ethics practices such as governance, conflicts of interest, informed consent, privacy, and use-and-access procedures, but that this information is often not as detailed as recommended in existing guidelines (AHRQ, 2020; EUnetHTA, 2019; Kalkman et al., 2019).

Regarding governance and conflicts of interest, Kalkman et al. (2019) stress that patient involvement and/or participation is crucial for a registry to develop and maintain public trust. However, the steering committees and/or governing boards of the registries in our sample rarely included patient representation. While some registries do provide patient participation initiatives separate from these formal bodies, it would be good if patients would also get a more formal role in deciding the policy of registries. Another point of improvement for registries would be to outline the individual responsibilities of all the members of their governing body instead of only outlining the responsibility of the body itself. Based on our results, this rarely happens, even though it is recommended by Chan et al. (2016) and features in the guidelines of both AHRQ (2020) and EUnetHTA (2019). Finally, while most registries provide information on how they are funded, only three also provided details about the influence these financiers have on the registry’s procedures. This leaves open the risk that external sponsors have unduly influence on the way the registry is governed, including potential impact on the scientific outcomes, something that has been widely labeled as undesirable (Dunn, Coiera, Mandl, & Bourgeois et al., 2016; Fabbri, Lai, Grundy & Bero, 2018; Lundh, Lexchin, Mitzes, Schroll, & Bero, 2017). Notably, this risk has been shown to be more pronounced for privately funded registries than for publicly funded registries (Jandhyala & Christopher, 2020).

Our finding that not all registries provided a consent form or patient information document is not necessarily problematic as some registries only provide anonymized data, which cannot be retraced to individuals and therefore does not require informed consent according to the European Union’s General Data Protection Regulation (GDPR). However, it must be noted that we based our coding on the information provided by the registries themselves. That is, if a registry stated that they use only anonymized data, we coded it as such in our dataset. The discussion around data anonymity is complex and it could be that registries misclassify the anonymization status of their data. If, for example, registries keep the patient data in a pseudonymized form (so patients can in principle be re-identified through their pseudonym), but release the data for use without the pseudonym, some experts in data safety might describe the release of the data as anonymized and others as pseudonymized. Registries that state that they only release anonymized data do not usually explain whether the data is also fully anonymized in the registry itself and if they do, they do not disclose what their anonymization procedure is.

If the data is not fully anonymized, it is essential to be clear about this when providing information as it involves a risk of identifying the patients. However, registries only rarely mentioned those risks when informing the patients. Other important elements like the aims of the registry were also not always provided even though this bit of information can be seen as the bare minimum in terms of informing current and potential registry participants (see Dyke & Hubbard, 2011; Rodriguez et al., 2009; World Medical Association, 2016). Registry websites typically also fail to provide information about if and how incidental findings will be communicated to patients, even though communicating incidental findings has far reaching ethical consequences and it is therefore important to include information and possibly an extra layer of consent in consent and information forms (Ells & Thombs, 2014; Saelaert, Mertes, Moerenhout, De Baere, & Devisch, 2020; Schaefer & Savulescu, 2018).

Finally, use-and-access procedures were often clearly described although it was sometimes hard to locate precisely where on the websites these procedures could be found. One point of attention for registries to focus on is to specify that data users should not try to re-identify the data they are provided. Even though this may be self-evident to some, it is seen as an important part of registry policies and thus important to emphasize (Knoppers, 2014; Nuffield Council on Bioethics, 2016).

In sum, we found that there is room for improvement in the way that patient registries provide information about ethics practices on their websites. But it is important to take these findings into context. For one, we investigated to what extent registry websites provide information about their procedures, but we did not assess the actual procedures themselves. It could, for example, be that a registry does have a detailed use-and-access policy but did not share this policy on their website. Providing procedural information to patients and researchers is important, especially with regard to ethics (AHRQ, 2020; EUnetHTA, 2019; Kalkman et al., 2019) but our findings should not necessarily be taken to mean that registries do not have ethics procedures in place, merely that they do not provide information about them.

Second, we focused on registries from European countries that were part of the ENCePP database. It could be that registries in other continents have better or worse information provision with regard to ethics practices or different procedures altogether. Moreover, during the data analysis for this study, the ENCePP website has been renewed and the database with patient registries has received an update. The database is now called the HMA-EMA Catalogue of real-world data sources, has improved its search functionality and now provides links between data sources and associated studies (European Network of Centres for Pharmacoepidemiology and Pharmacovigilance, 2024). The new database (see https://catalogues.ema.europa.eu/catalogue-rwd-sources) also involves updates of all the patient registries, so future studies like ours will have access to more recent data, hopefully simplifying the search process.

An interesting venue for future research would be the difference between commercial and non-commercial registries. Based on our experience in this study, large commercial registries typically have elaborate websites but fail to provide many bits of relevant information, while non-commercial registries have smaller websites but provide more relevant information. However, we did not have sufficient data to do a rigorous evaluation so future studies may look into this difference in information denseness more systematically. Aside from ethics practices, we believe the effect of commercial funding on patient registries is an interesting area of research in and of itself given that the literature so far has focused more on the difference between commercial and non-commercial clinical trials (e.g., Camps, Rodríguez, and Agustí, 2018; Nejstgaard, Laursen, Lundh, and Hróbjartsson, 2023; Schott et al., 2010). One could for example check whether commercially funded registries provide less or more guidance with regard to research transparency (i.e., whether they mandate studies to be preregistered or results to be shared), and whether studies from commercial registries less often or more often contain statistically significant results.

A secondary benefit of our study could be the checklist that we developed to assess the patient registries. We believe that it captures the most important ethics practices relevant to patient registries and provides a relatively simple way to assess these practices. The owners of patient registries could use the checklist to see where they can improve their information provision, and meta-researchers could use the checklist to find general trends in how patient registries communicate their ethical procedures. One thing to take into consideration when using this checklist is that it is based on three sources that we deemed pertinent. As such, it is not based on a systematic review of the literature. To illustrate that, an additional source that we did not include is the Guideline on registry-based studies by the European Medicines Agency (2021). We chose not to include this guideline because we felt that it did not add any ethics elements that we did not already capture by using the other three sources. This assessment is supported by the fact that the main ethics themes in the EMA guideline (governance, informed consent and data protection, and funding) are included in our checklist. That being said, we encourage registry owners, researchers, clinicians, and patients to identify any missing or irrelevant elements in our checklist and tweak the checklist to fit their needs.

## Data Availability

All data is available at https://osf.io/wmrv2

## Declarations

### Ethics approval and consent to participate

Not applicable.

### Consent for publication

Not applicable.

### Availability of data and materials

All data and materials used in this study can be found in the Open Science Framework repository of the project: https://osf.io/4vh5q.

### Funding

We have received funding from the European Union’s Horizon Europe Research and Innovation Actions under grant no. 101095479 (More-EUROPA). Views and opinions expressed are however those of the authors only and do not necessarily reflect those of the European Union nor the granting authority.

Neither the European Union nor the granting authority can be held responsible for them.

### Authors’ contributions

Conceptualization: DS

Methodology: OvdA, SS & DS

Investigation: OvdA

Data curation: OvdA

Writing – Original Draft: OvdA

Writing – Review & Editing: SS & DS

Supervision: DS

Funding acquisition: DS

## Acknowledgements

Not applicable.

## References

Agency for Healthcare Research and Quality. Registries for Evaluating Patient Outcomes: A User’s Guide. 4th ed. 2020.

Chan T, Di Iorio CT, De Lusignan S, Russo DL, Kuziemsky C, Liaw ST. UK National Data Guardian for Health and Care. J Innov Health Inform. 2016;23(3). DOI: 10.14236/jhi.v23i3.818

Dunn AG, Coiera E, Mandl KD, Bourgeois FT. Conflict of interest disclosure in biomedical research: a review of current practices, biases, and the role of public registries in improving transparency. Res Integr Peer Rev. 2016;1:1–8. DOI: 10.1186/s41073-016-0007-6

Dyke SO, Hubbard TJ. Developing and implementing an institute-wide data sharing policy. Genome Med. 2011;3(9):1–8. DOI: 10.1186/gm215

Ells C, Thombs BD. The ethics of how to manage incidental findings. CMAJ. 2014;186(9):655–656. DOI: 10.1503/cmaj.130950

European Network of Centres for Pharmacoepidemiology and Pharmacovigilance. Launch of the HMA-EMA Real-World Data (RWD) Catalogues and New ENCePP Website. https://encepp.europa.eu/newsroom/news/launch-hma-ema-real-world-data-rwd-catalogues-and-new-encepp-website-2024-02-15_en. Published February 15, 2024. Accessed April 10, 2024.

European Network for Health Technology Assessment. Registry Evaluation and Quality Standards Tool (REQueST). 2019. Available from: https://www.eunethta.eu/request-tool-and-its-vision-paper

Fabbri A, Lai A, Grundy Q, Bero LA. The influence of industry sponsorship on the research agenda: a scoping review. Am J Public Health. 2018;108(11):e9–e16. DOI: 10.2105/AJPH.2018.304586

Camps I, Rodríguez A, Agustí A. Non-commercial vs. commercial clinical trials: a retrospective study of the applications submitted to a research ethics committee. Br J Clin Pharmacol. 2018;84(6):1384–1388. DOI: 10.1111/bcp.13576

European Medicines Agency. Guideline on registry-based studies. https://www.ema.europa.eu/en/documents/scientific-guideline/guideline-registry-based-studies_en.pdf-0. Published October 22, 2021. Accessed April 10, 2024.

Jandhyala R, Christopher S. Factors influencing the generation of evidence from simple data held in international rare disease patient registries. Pharm Med. 2020;34(1):31–38. DOI: 10.1007/s40290-019-00338-7

Kalkman S, Mostert M, Gerlinger C, van Delden JJ, van Thiel GJ. Responsible data sharing in international health research: a systematic review of principles and norms. BMC Med Ethics. 2019;20:1–13. DOI: 10.1186/s12910-019-0416-x

Knoppers BM. Framework for responsible sharing of genomic and health-related data. HUGO J. 2014;8(1):3. DOI: 10.1186/s11568-014-0003-1

Langhof H, Schwietering J, Strech D. Practice evaluation of biobank ethics and governance: current needs and future perspectives. J Med Genet. 2018. doi:10.1136/jmedgenet-2018-105617

Lundh A, Lexchin J, Mintzes B, Schroll JB, Bero L. Industry sponsorship and research outcome. Cochrane Database Syst Rev. 2017;(2). DOI: 10.1002/14651858.MR000033.pub3

National Commission for the Protection of Human Subjects of Biomedical and Behavioral Research. The Belmont report: Ethical principles and guidelines for the protection of human subjects of research. U.S. Department of Health and Human Services. 1979. Available from: https://www.hhs.gov/ohrp/regulations-and-policy/belmont-report/read-the-belmont-report/index.html

Nejstgaard CH, Laursen DRT, Lundh A, Hróbjartsson A. Commercial funding and estimated intervention effects in randomized clinical trials: Systematic review of meta-epidemiological studies. Res Synth Methods. 2023;14(2):144–155. DOI: 10.1002/jrsm.1500

Nuffield Council on Bioethics. The collection, linking and use of data in biomedical research and health care: ethical issues. 2015. Available from: https://www.nuffieldbioethics.org/publications/biological-and-health-data

Pavlenko E, Strech D, Langhof H. Implementation of data access and use procedures in clinical data warehouses. A systematic review of literature and publicly available policies. BMC Med Inform Decis Mak. 2020;20(1):157. DOI: 10.1186/s12911-020-01177-z

Plueschke K, Jonker C, Strassmann V, Kurz X. Collection of Data on Adverse Events Related to Medicinal Products: A Survey Among Registries in the ENCePP Resources Database. Drug Saf. 2022;45(7):747–754. DOI: 10.1007/s40264-022-01172-3

Rodriguez H, Snyder M, Uhlén M, Andrews P, Beavis R, Borchers C, et al. Recommendations from the 2008 international summit on proteomics data release and sharing policy: The Amsterdam principles. J Proteome Res. 2009;8(7):3689–3692. DOI: 10.1021/pr900023v

Saelaert M, Mertes H, Moerenhout T, De Baere E, Devisch I. Ethical values supporting the disclosure of incidental and secondary findings in clinical genomic testing: a qualitative study. BMC Med Ethics. 2020;21:1–12. DOI: 10.1186/s12910-020-00492-7

Schaefer GO, Savulescu J. The right to know: a revised standard for reporting incidental findings. Hastings Cent Rep. 2018;48(2):22–32. DOI: 10.1002/hast.813

Schott G, Pachl H, Limbach U, Gundert-Remy U, Ludwig WD, Lieb K. The financing of drug trials by pharmaceutical companies and its consequences: Part 1: a qualitative, systematic review of the literature on possible influences on the findings, protocols, and quality of drug trials. Dtsch Arztebl Int. 2010;107(16):279. DOI: 10.3238/arztebl.2010.0279

Schwietering J, Langhof H, Strech D. Empirical studies on how ethical recommendations are translated into practice: a cross-section study on scope and study objectives. BMC Med Ethics. 2023;24(1):2. DOI: 10.1186/s12910

World Medical Association, WMA. Declaration of Taipei on Ethical Considerations Regarding Health Databases and Biobanks. 2016.

